# The landscape of host genetic factors involved in immune response to common viral infections

**DOI:** 10.1101/2020.05.01.20088054

**Authors:** Linda Kachuri, Stephen S. Francis, Maike Morrison, George A. Wendt, Yohan Bossé, Taylor B. Cavazos, Sara R. Rashkin, Elad Ziv, John S. Witte

## Abstract

**Introduction:** Humans and viruses have co-evolved for millennia resulting in a complex host genetic architecture. Understanding the genetic mechanisms of immune response to viral infection provides insight into disease etiology and therapeutic opportunities.

**Methods:** We conducted a comprehensive study including genome-wide and transcriptome-wide association analyses to identify genetic loci associated with immunoglobulin G antibody response to 28 antigens for 16 viruses using serological data from 7924 European ancestry participants in the UK Biobank cohort.

**Results:** Signals in human leukocyte antigen (HLA) class II region dominated the landscape of viral antibody response, with 40 independent loci and 14 independent classical alleles, 7 of which exhibited pleiotropic effects across viral families. We identified specific amino acid (AA) residues that are associated with seroreactivity, the strongest associations presented in a range of AA positions within DRβi at positions 11, 13, 71, and 74 for Epstein-Barr Virus (EBV), Varicella Zoster Virus (VZV), Human Herpes virus 7, (HHV7) and Merkel cell polyomavirus (MCV). Genome-wide association analyses discovered 7 novel genetic loci outside the HLA associated with viral antibody response (*P*<5.0×10^-8^), including *FUT2* (19q13.33) for human polyomavirus BK (BKV), *STING1* (5q31.2) for MCV, as well as *CXCR5* (11q23.3) and *TBKBP1* (17q21.32) for HHV7. Transcriptome-wide association analyses identified 114 genes associated with response to viral infection, 12 outside of the HLA region, including *ECSCR*: P=5.0*10^-15^ (MCV), *NTN5:* P=1.1×10^-9^ (BKV), and *P2RY13:* P=1.1×10^-8^ EBV nuclear antigen. We also demonstrated pleiotropy between viral response genes and complex diseases; from autoimmune disorders to cancer to neurodegenerative and psychiatric conditions.

**Conclusions:** Our study confirms the importance of the HLA region in host response to viral infection and elucidates novel genetic determinants beyond the HLA that contribute to host-virus interaction.

## INTRODUCTION

Viruses have been infecting cells for a half a billion years^1^. During our extensive co-evolution viruses have exerted significant selective pressure on humans and vice versa; overtly during fatal outbreaks, and covertly through cryptic immune interaction when a pathogen remains latent. The recent pandemic of severe acute respiratory syndrome coronavirus 2 (SARS-CoV-2) highlights the paramount public health need to understand human genetic variation in response to viral challenge. Clinical variation in COVID-19 severity and symptomatic presentation may be due to differences host genetic factors relating to immune response^2^. Furthermore, many common infections are cryptically associated with a variety of complex illnesses, especially those with an immunologic component, from cancer to autoimmune and neurologic conditions^3-5^. Despite their broad health relevance, few large-scale genome-wide association studies (GWAS) have been conducted on serological response phenotypes^6-10^. Understanding the genetic architecture of immunologic response to viruses may therefore provide new insight into etiologic mechanisms of diverse complex diseases.

Several common viruses exert a robust cell mediated and humoral immune response that bi-directionally modulate the balance between latent and lytic infection. Studies have demonstrated a strong heritable component (32-48%) of antibody response^11^ and identified associations between host polymorphisms in genes relating to cell entry, cytokine production, and immune response and a variety of viruses^12^. The predominance of previously reported associations with have implicated genetic variants in human leucocyte antigen (HLA) class I and II genes in the modulation of immune response to diverse viral antigens^7,13^.

In this study we utilize data from the UK Biobank (UKB) cohort^14^ to evaluate the relationship between host genetics and immunoglobulin G antibody response to 28 antigens for 16 viruses. Immunoglobulin G (IgG) antibody is the most common antibody in blood, which serves as a stable biomarker of lifetime exposure to common viruses. High levels of specific IgG’s can be the result of chronic infection, while low levels may indicate poor immunity. Viruses assayed in the UKB multiplex serology panel were previously chosen based on putative links to chronic diseases including cancer, autoimmune, and neurodegenerative conditions^15^. We conduct integrative genome-wide and transcriptome-wide analyses of antibody response and positivity to viral antigens (**Figure 1**), which elucidate novel genetic underpinnings of viral infection and immune response.

**Figure 1:**
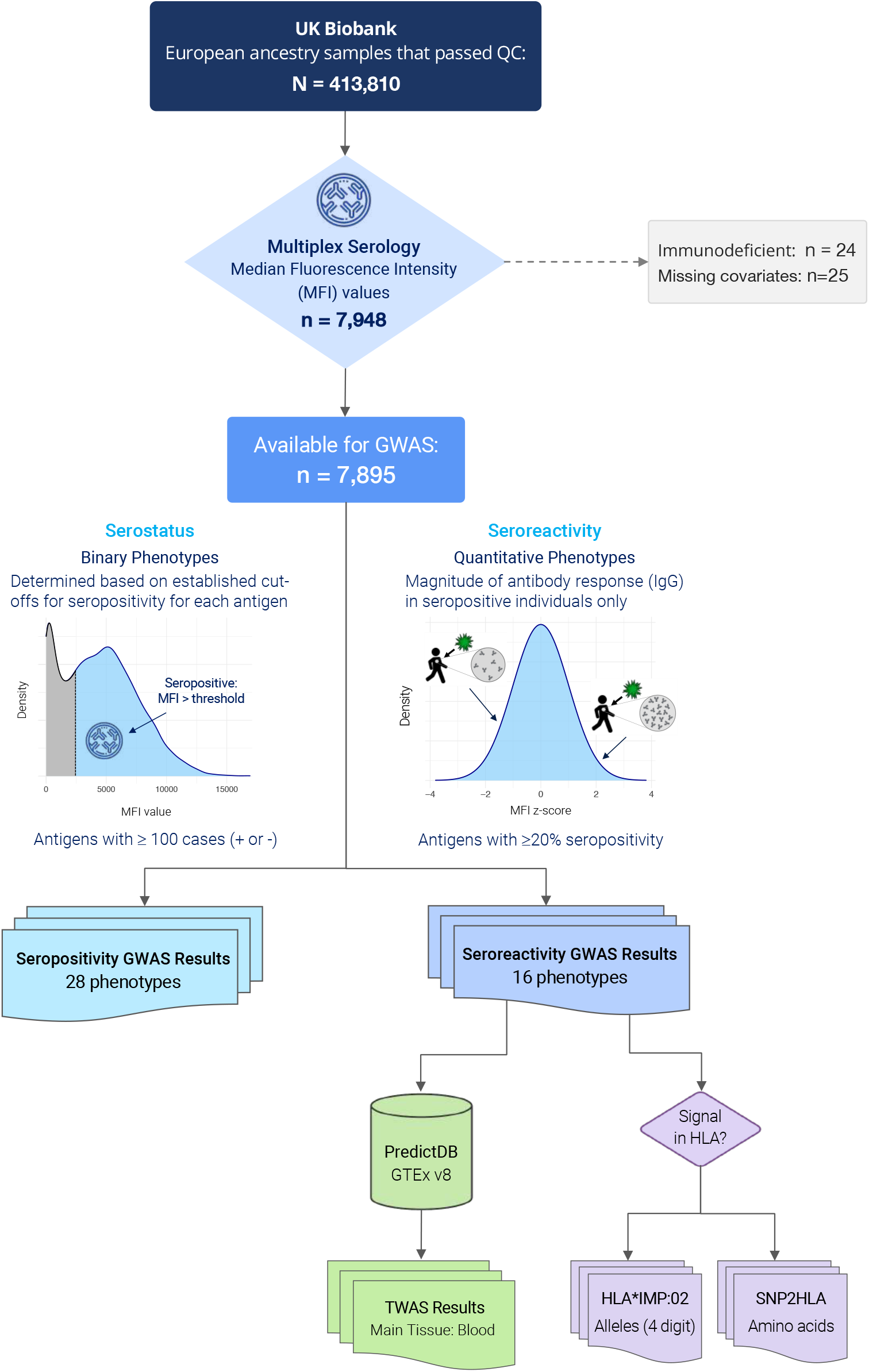
Flow chart describing the main serological phenotypes and association analyses

## METHODS

### Study Population and Phenotypes

The UK Biobank (UKB) is a population-based prospective cohort of over 500,000 individuals aged 40-69 years at enrollment in 2006-2010 who completed extensive questionnaires, physical assessments, and provided blood samples^14^. Analyses were restricted to individuals of predominantly European ancestry based on self-report and after excluding samples with any of the first two genetic ancestry principal components (PCs) outside of 5 standard deviations (SD) of the population mean (**Supplementary Figure 1**). We removed samples with discordant self-reported and genetic sex, samples with call rates <97% or heterozygosity >5 SD from the mean, and one sample from each pair of first-degree relatives identified using KING^16^.

Of the 413,810 European ancestry individuals available for analysis, a total of 7948 had serological measures. A multiplex serology panel (IgG) was performed over a 2-week period using previously developed methods^17,18^ that have been successfully applied in epidemiological studies^7,19^. Details of the serology methods and assay validation performance are described in Mentzer et al.^15^ Briefly, multiplex serology was performed using a bead-based glutathione S-transferase (GST) capture assay with glutathione-casein coated fluorescence-labelled polystyrene beads and pathogen-specific GST-X-tag fusion proteins as antigens^15^. Each antigen was loaded onto a distinct bead set and the beads were simultaneously presented to primary serum antibodies at serum dilution 1: 1000^15^. Immunocomplexes were quantified using a Luminex 200 flow cytometer, which produced Median Fluorescence Intensities (MFI) for each antigen. The serology assay showed adequate performance, with a median coefficient of variation (CV) of 17% across all antigens and 3.5% among seropositive samples only^15^.

### Genome-Wide Association Analysis

We evaluated the relationship between genetic variants across the genome and serological phenotypes using PLINK 2.0 (October 2017 version). Participants were genotyped on the Affymetrix Axiom UK Biobank array (89%) or the UK BiLEVE array (11%)^14^ with genome-wide imputation performed using the Haplotype Reference Consortium data and the merged UK10K and 1000 Genomes phase 3 reference panels^14^. We excluded variants out of Hardy-Weinberg equilibrium at p<1×10^-5^, call rate <95% (alternate allele dosage within 0.1 of the nearest hard call to be non-missing), imputation quality INFO<0.30, and MAF<0.01.

Seropositivity for each antigen was determined using established cut-offs based on prior validation work^15^. The primary GWAS focused on continuous phenotypes (MFI values), which measure the magnitude of antibody response, also referred to as seroreactivity. These analyses were conducted among seropositive individuals only for antigens with seroprevalence of ≥20% (n=1500) based on 80% power to detect only common variants with large effect sizes at this sample size (**Supplementary Figure 2**). MFI values were transformed to standardized, normally distributed z-scores using ordered quantile normalization^20^.

**Figure 2:**
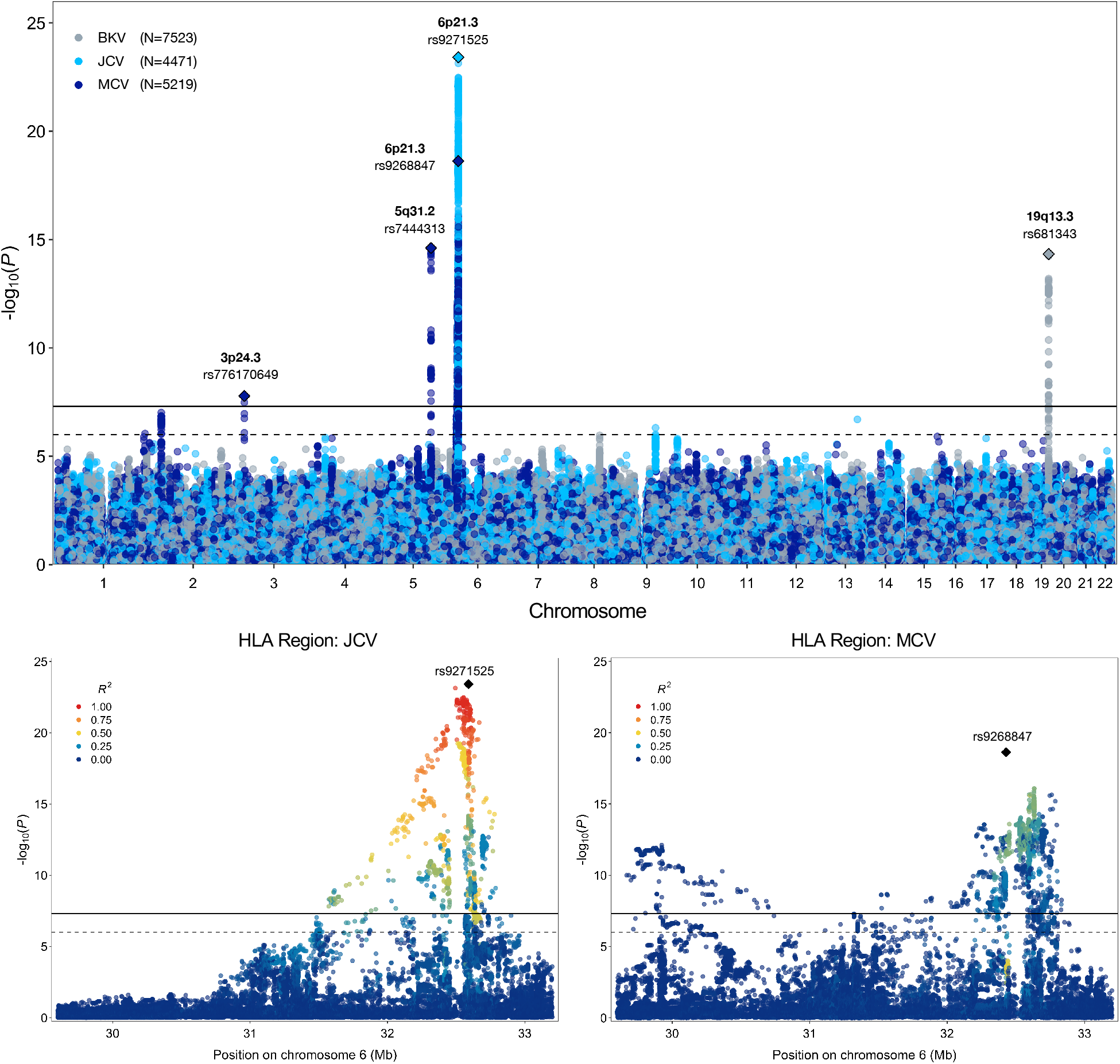
Results from genome-wide and regional association analyses of continuous antibody response phenotypes (MFI z-scores) among individuals seropositive for human polyomaviruses BKV, JCV, and Merkel cell (MCV). The lower two panels depict the association signal and linkage disequilibrium (LD) structure in the HLA region for JCV and MCV.

Seroreactivity GWAS was conducted using linear regression with adjustment for age at enrollment, sex, body-mass index (BMI), socioeconomic status (Townsend deprivation index), the presence of any autoimmune and/or inflammatory conditions, genotyping array, serology assay date, quality control flag indicating sample spillover or an extra freeze/thaw cycle, and the top 10 genetic ancestry principal components (PC’s). Autoimmune and chronic inflammatory conditions were identified using the following primary and secondary diagnostic ICD-10 codes (E10, M00-03, M05-M14, M32, L20-L30, L40, G35, K50-52, K58, G61) in Hospital Episode Statistics. Individuals diagnosed with any immunodeficiency (ICD-10 D80-89, n=24) were excluded from all analyses.

For all antigens with at least 100 seropositive (or seronegative for pathogens with ubiquitous exposure) individuals, GWAS of discrete seropositivity phenotypes was undertaken using logistic regression, adjusting for the same covariates listed above.

The functional relevance of the lead GWAS loci for antibody response was assessed using in-silico functional annotation analyses based on Combined Annotation Dependent Depletion (CADD)^21^ scores and RegulomeDB 2.0^22^, and by leveraging external datasets, such as GTEx v8, DICE (Database of Immune Cell Expression)^23^, and the Human Plasma Proteome Atlas^24,25^.

### Cross-Trait Associations with Disease

We explored pleiotropic associations between lead variants influencing antibody levels and several chronic diseases with known or hypothesized viral risk factors. Associations with selected cancers were obtained from a cancer pleiotropy meta-analysis of the UK Biobank and Genetic Epidemiology Research on Aging cohorts^26^. Summary statistics for the schizophrenia GWAS of 33,640 cases and 43,456 controls by Lam et al.^27^ were downloaded from the Psychiatric Genomics Consortium. Association p-values were obtained from the National Institute on Aging Genetics of Alzheimer’s Disease Data Storage Site for the GWAS by Jun et al.^28^, which included 17,536 cases and 53,711 controls. Associations with p<7.3×10^-4^ were considered statistically significant after correction for the number of variants and phenotypes tested.

### HLA Regional Analysis

For phenotypes displaying a genome-wide significant signal in the HLA region, independent association signals were ascertained using two complementary approaches: clumping and conditional analysis. Clumping is a post-processing step applied to GWAS summary statistics to identify independent association signals by grouping variants based on LD within specific windows. Clumping was performed on all variants with *P*<5×10^-8^ for each phenotype, as well as across phenotypes. Clumps were formed around index variants with the lowest p-value and all other variants with LD *r*^2^>0.05 within a ±500 kb window were considered non-independent and assigned to that variant’s clump.

Next, we conducted conditional analyses using a forward stepwise strategy to identify statistically independent signals within each type of variant (SNP/indel or classical HLA allele). Unlike clumping, conditional analyses involve fitting a new model that includes specific variants as covariates, thereby directly accounting for LD and providing association estimates that are adjusted for other relevant SNP effects. A total of 38,655 SNPs/indels on chromosome 6 (29,600,000 – 33,200,000 bp) were extracted to conduct regional analyses. Classical HLA alleles were imputed for UKB participants at 4-digit resolution using the HLA*IMP:02 algorithm^14^, with modified settings to accommodate the addition of diverse samples from population reference panels described by Motyer et al.^29^. Details of the HLA imputation procedure are described in UKB Resource 182. Imputed dosages were available for 362 classical alleles in 11 genes: *HLA-A, HLA-B*, and *HLA-C* (class I); *HLA-DRB5, HLA-DRB4, HLA-DRB3, HLA-DRB1, HLA-DQA1, HLA-DQB1, HLA-DPA1*, and *HLA-DPB1* (class II). Allele names with “99:01” for DRB3/4/5, which denote copy number absence, were renamed as “00:00” to avoid confusion with traditional HLA nomenclature. We also used SNP2HLA^30^ to impute HLA alleles and corresponding amino acid sequences at a four-digit resolution in *HLA-A, HLA-B, HLA-C, HLA-DRB1, HLA-DQA1, HLA-DQB1, HLA-DPA1* and *HLA-DPB1* using the Type 1 Diabetes Genetics Consortium (T1DGC) reference panel comprised of 2,767 unrelated individuals of European descent. T1DGC was also among several reference datasets used by HLA*IMP:02.

Analyses were restricted to common HLA alleles and amino acid sequences (frequency ≥ 0.01) with imputation quality scores >0.30, for a total of 1081 markers (101 alleles + 980 amino acid residues). Linear regression models were adjusted for the same set of covariates as the GWAS. Associations for each marker were considered statistically significant if *P*<4.6×10^-5^ based on Bonferroni correction for 1081 tests.

For each antigen response phenotype, we identified SNPs/indels or classical HLA alleles with the lowest p-value, among variants that achieved Bonferroni-significant associations (*P*<4.6×10^-5^), and performed forward iterative conditional regression to identify other independent signals, until no associations with a conditional p-value (*P*_cond_)<5×10^-8^ remained. We also assessed the independence of associations across different types of genetic variants by including conditionally independent HLA alleles as covariates in the SNP-based analysis.

For amino acid positions with >2 possible residues (alleles), we applied the haplotype omnibus test to obtain an overall p-value for jointly testing all possible substitutions at that specific position. The omnibus test was applied to all amino acid residues at a given position, even if not all substitutions achieved the Bonferroni-corrected threshold (*P*<4.6×10^-5^) in the single-marker analysis. The frequency of amino acid substitutions at specific HLA alleles was determined using European ancestry reference populations part of the Allele Frequency Net Database (AFND 2020)^31^.

### Transcriptome-Wide Association Analysis

Gene transcription levels were imputed and analyzed using the MetaXcan approach^32^, applied to GWAS summary statistics for quantitative antigen phenotypes. For imputation, we used biologically informed MASHR-M prediction models^33^ based on GTEx v8 with effect sizes computed using MASHR (Multivariate Adaptive Shrinkage in R)^34^ for variants fine-mapped with DAP-G (Deterministic Approximation of Posteriors)^35,36^. An advantage of this approach is that MASHR effect sizes are smoothed by taking advantage of the correlation in cis-eQTL effects across tissues. For each antigen, we performed a transcriptome-wide association study (TWAS) using gene expression levels in whole blood. Statistically significant associations for each gene were determined based on Bonferroni correction for the number of genes tested.

We also examined gene expression profiles in tissues that represent known infection targets or related pathologies. Human herpesviruses and polyomaviruses are neurotropic and have been implicated in several neurological conditions^37,38^, therefore we considered gene expression in the frontal cortex. For Epstein-Barr virus (EBV) antigens additional models included EBV-transformed lymphocytes. Merkel cell polyomavirus (MCV) is a known cause of Merkel cell carcinoma^39^, a rare but aggressive type of skin cancer, therefore we examined transcriptomic profiles in skin tissues for MCV only.

Pathways represented by genes associated with antibody response to viral antigens were summarized by conducting enrichment analysis using curated Reactome gene sets and by examining protein interaction networks using the STRING database^40^. Significantly associated TWAS genes were grouped by virus family (herpesviruses vs. polyomaviruses) and specificity of association (multiple antigens vs. single antigen).

## RESULTS

A random sample of the participants representative of the full UKB cohort was assayed using a multiplex serology panel^15^. We analyzed data from 7924 participants of predominantly European ancestry, described in **Supplementary Table 1**. Approximately 90% of individuals were seropositive for herpes family viruses with ubiquitous exposure: EBV (EBV EA-D: 86.2% to ZEBRA: 91.2%), Human Herpesvirus 7 (HHV7 94.8%), and Varicella Zoster Virus (VZV 92.3%). Seroprevalence was somewhat lower for cytomegalovirus (CMV), ranging between 56.5% (CMV pp28) and 63.3% (CMV pp52), and Herpes Simplex virus-1 (HSV1 69.3%). Human polyomavirus BKV was more prevalent (95.3%) compared to other polyomaviruses, Merkel cell polyoma virus (MCV 66.1%) and polyomavirus JC (JCV) (56.6%). Less common infections included HSV-2 (15.2%), HPV16 (E6 and E7 oncoproteins: 4.7%), HPV18 (2.4%), Human T-cell lymphotropic virus type 1 (HTLV1, 1.6%), Hepatitis B (HBV, 1.6%), and Hepatitis C (HCV, 0.3%).

### Genetic Determinants of Response to Viral Infection

Results from our GWAS of antibody response phenotypes were dominated by signals in the HLA region, which were detected for all EBV antigens (EA-D, EBNA, p18, ZEBRA), CMV pp52, HSV1, HHV7, VZV, JCV and MCV (**Table 1; Supplementary Figure 3**). Most of the top-ranking HLA variants for each antigen were independent of those for other antigens based on *r*^2^ but not D’ (**Supplementary Figure 4**). Exceptions were moderate LD between lead variants for EBV ZEBRA and HSV1 (*r*^2^=0.45), EBV EBNA and JCV (*r*^2^=0.45), and HHV7 and MCV (*r*^2^=0.44). However, based on the complex LD structure and effect sizes, we cannot rule out that these linked to rare haplotypes. Outside of the HLA region, genome-wide significant associations with seroreactivity were detected for: MCV at 3p24.3 (rs776170649, *LOC339862: P*=1.7×10^-^8) and 5q31.2 (rs7444313, *TMEM173* (also known as *STING1): P*=2.4×10^-15^); BKV at 19q13.3 (rs681343, *FUT2: P*=4.7×10^-15^) (**Figure 2**); EBV EBNA at 3q25.1 (rs67886110, *MED12L: P*=1.3×10^-9^); HHV-7 at 11q23.3 (rs75438046, *CXCR5: P*=1.3×10^-8^) and 17q21.3 (rs1808192, *TBKBP1*: P=9.8×10^-9^); and HSV-1 at 10q23.3 (rs11203123: *P*=3.9×10^-8^). However, the loci outside of HLA identified for HHV7 and HSV1 were not statistically significant considering a more stringent significance threshold corrected for the number of seroreactivity phenotypes tested (*P* < 5.0×10^-8^/16 = 3.1×10^-9^).

**Table 1:**
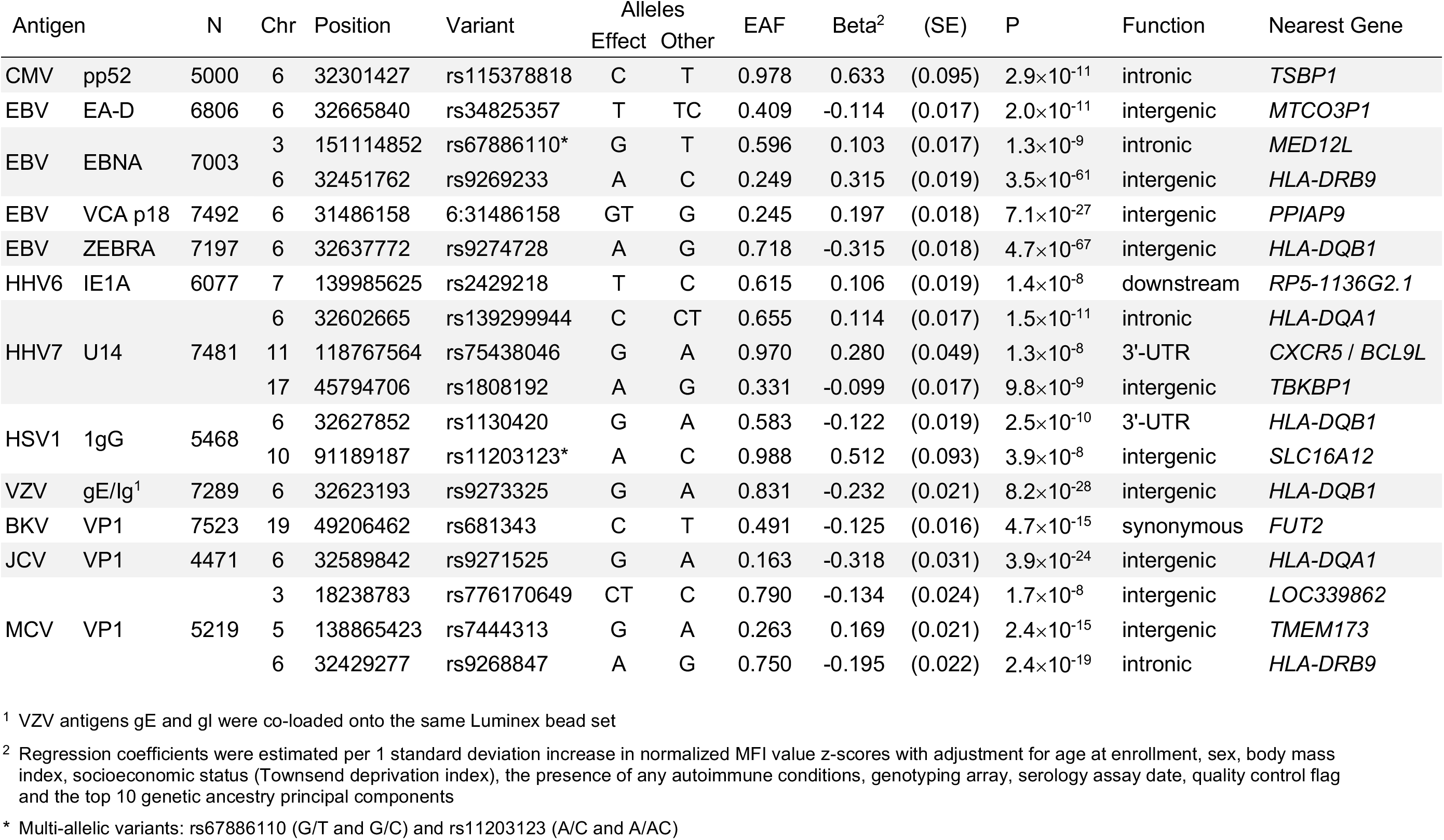
Lead genome-wide significant variants (*P*<5.0×10^-8^) for continuous antibody response phenotypes for antigens with at least 20% seroprevalence.

GWAS of discrete seropositivity phenotypes identified associations in HLA for EBV EA-D (rs2395192: OR=0.66, *P*=4.0×10^-19^), EBV EBNA (rs9268848: OR=1.60, *P*=1.2×10^-18^), EBV ZEBRA (rs17211342: 0.63, *P*=1.6×10^-15^), VZV (rs3096688: OR=0.70, *P*=3.7×10^-8^), JCV (rs9271147: OR=0.54, *P*=1.3×10^-42^), and MCV (rs17613347: OR=0.61, *P*=1.2×10^-26^) (**Supplementary Figure 2; Supplementary Table 3**). An association with susceptibility to MCV infection was also observed at 5q31.2 (rs1193730215, *ECSCR:* OR=1.26, *P*=7.2×10^-9^), with high LD (*r*^2^=0.95) between seroreactivity and seropositivity lead variants.

Several genome-wide significant associations were observed for antigens with <20% seroprevalence, which were not included in the GWAS of antibody response due to inadequate sample size (**Supplementary Table 3**). Infection susceptibility variants were identified for HSV2 in 17p13.2 (rs2116443: OR=1.28, *P*=4.5×10^-8^; *ITGAE*); HPV16 E6 and E7 oncoproteins in 6p21.32 (rs601148: OR=0.60, *P*=3.3×10^-9^; *HLA-DRB1)* and 19q12 (rs144341759: OR=0.383, *P*=4.0×10^-8^; *CTC-448F2.6*); and HPV18 in 14q24.3 (rs4243652: OR=3.13, *P*=7.0×10^-10^). Associations were also detected for Kaposi’s sarcoma-associated herpesvirus (KSHV), HTLV1, HBV and HCV, including a variant in the *MERTK* oncogene (HCV Core rs199913364: OR=0.25, *P*=1.2×10^-8^). After correcting for 28 serostatus phenotypes tested (*P*<1.8×10^-9^), the only statistically significant associations remained for EBV EA-D (rs2395192), EBV EBNA (rs9268848), EBV ZEBRA (rs17211342), JCV (rs9271147), MCV (rs17613347), and HPV18 (rs4243652).

### Functional Characterization of GWAS Findings

In-silico functional analyses of the lead 17 GWAS variants identified enrichment for multiple regulatory elements (summarized in **Supplementary Table 4**). Three variants were predicted to be in the top 10% of deleterious substitutions in GRCh37 based on CADD scores >10: rs776170649 (MCV, CADD=15.61), rs139299944 (HHV7, CADD=12.15), and rs9271525 (JCV, CADD=10.73). Another HHV7-associated variant, rs1808192 (RegulomeDB rank: 1f), an eQTL and sQTL for *TBKBP1*, mapped to 44 functional elements for multiple transcription factors, including IKZF1, a critical regulator of lymphoid differentiation frequently mutated in B-cell malignancies.

Eleven sentinel variants were eQTLs and 8 were splicing QTLs in GTEx, with significant (FDR<0.05) effects across multiple genes and tissues (**Supplementary Figure 5**). The most common eQTL and sQTL targets included *HLA-DQA1, HLA-DQA2, HLA-DQB1, HLA-DQB2, HLA-DRB1*, and *HLA-DRB6*. Outside of HLA, rs681343 (BKV), a synonymous *FUT2* variant was an eQTL for 8 genes, including *FUT2* and *NTN5*. MCV variant in 5q31.2, rs7444313, was an eQTL for 7 genes, with concurrent sQTL effects on *TMEM173*, also known as *STING1* (stimulator of interferon response cGAMP interactor 1) and *CXXC5*. Gene expression profiles in immune cell populations from DICE^23^ identified several cell-type specific effects that were not observed in GTEx. An association with *HLA-DQB1* expression in CD4+ T_H_2 cells was observed for rs9273325, 6:31486158_GT_G was an eQTL for *ATP6V1G2* in naïve CD4+ T cells, and rs1130420 influenced the expression of 8 HLA class II genes in naïve B-cells and CD4+ T_H_17 cells.

We identified 7 significant (p<5.0×10^-8^) protein quantitative trait loci (pQTL) for 38 proteins (**Supplementary Table 5**). Most of the pQTL targets were components of the adaptive immune response, such as the complement system (C4, CFB), chemokines (CCL15, CCL25), and defensin processing (Beta-defensin 19, Trypsin-3). The greatest number and diversity of pQTL targets (n=16) was observed for rs681343, including BPIFB1, which plays a role in antimicrobial response in oral and nasal mucosa^41^; FUT3, which catalyzes the last step of Lewis antigen biosynthesis; and FGF19, part of the PI3K/Akt/MAPK signaling cascade that is dysregulated in cancer and neurodegenerative diseases^42^.

### Cross-trait associations with disease outcomes

To contextualize the relevance of genetic loci involved in infection response, we explored associations with selected cancers, schizophrenia, and that have a known or suspected viral etiology (**Supplementary Table 6**). The strongest secondary signal was observed for rs9273325 (*HLA-DQB1*), which was negatively associated with VZV antibody response and positively associated with schizophrenia susceptibility (OR=1.13, *P*=4.3×10^-15^). Other significant (Bonferroni *P*<7.4×10^-4^) associations with schizophrenia were detected for HSV1 (rs1130420: OR=1.06, *P*=1.8×10^-5^), EBV EA-D (rs2647006: OR=0.96, *P*=2.7×10^-4^), JCV (rs9271525: OR=1.06, *P*=6.8×10^-5^) and BKV (rs681343: OR=0.96, *P*=2.5×10^-4^), with the latter being the only pleiotropic signal outside of HLA. Inverse associations with hematologic cancers were observed for HSV1 (rs1130420: OR=0.89, *P*=3.5×10^-6^), VZV (rs9273325: OR=0.88, *P*=4.4×10^-5^), and EBV EBNA (rs9269233: OR=0.88, *P*=2.7×10^-4^) variants. HSV1 antibody response was also linked to Alzheimer’s disease (rs1130420: *P*=1.2×10^-4^).

### Regional HLA Associations

Associations within the HLA region were refined by identifying independent (LD *r*^2^<0.05 within ±500kb) index variants with *P*<5.0×10^-8^ for each antigen response phenotype (**Supplementary Table 7**). Clumping seropositivity associations with respect to lead antibody response variants did not retain any loci, suggesting non-independence in signals for infection and reactivity for the same antigen. For this reason, all subsequent analyses focus on seroreactivity phenotypes. Clumping across phenotypes to assess the independence of HLA associations for different antigens identified 40 independent index variants: EBV EBNA (12), VZV (11), EBV ZEBRA (8), EBV p18 (5), MCV (3), and EBV EA-D (1) (**Supplementary Table 9**). No LD clumps were anchored by variants detected for CMV pp52, HHV7, HSV1, or JCV, suggesting that the HLA signals for these antigens are captured by lead loci for other phenotypes. The largest region with the lowest p-value was anchored by rs9274728 (*P*=4.7×10^-67^) near *HLA-DQB1*, originally detected for EBV ZEBRA. Of the 11 VZV-associated variants, the largest clump was formed around rs4990036 (*P*=4.5×10^-26^) in *HLA-B*.

Iterative conditional analyses adjusting for the HLA SNP/indel with the lowest p-value were performed until no variants remained with *P*_cond_<5.0×10^-8^. Additional independent variants were identified for EBV EBNA (rs139299944, rs6457711, rs9273358, rs28414666, rs3097671), EBV ZEBRA (rs2904758, rs35683320, rs1383258), EBV p18 (rs6917363, rs9271325, rs66479476), and MCV (rs148584120, rs4148874) (**Figure 3; Supplementary Table 8**). For CMV pp52, HHV7, HSV1, JCV, and VZV, the regional HLA signal was captured by the top GWAS variant (**Figure 2; Supplementary Table 8**).

**Figure 3:**
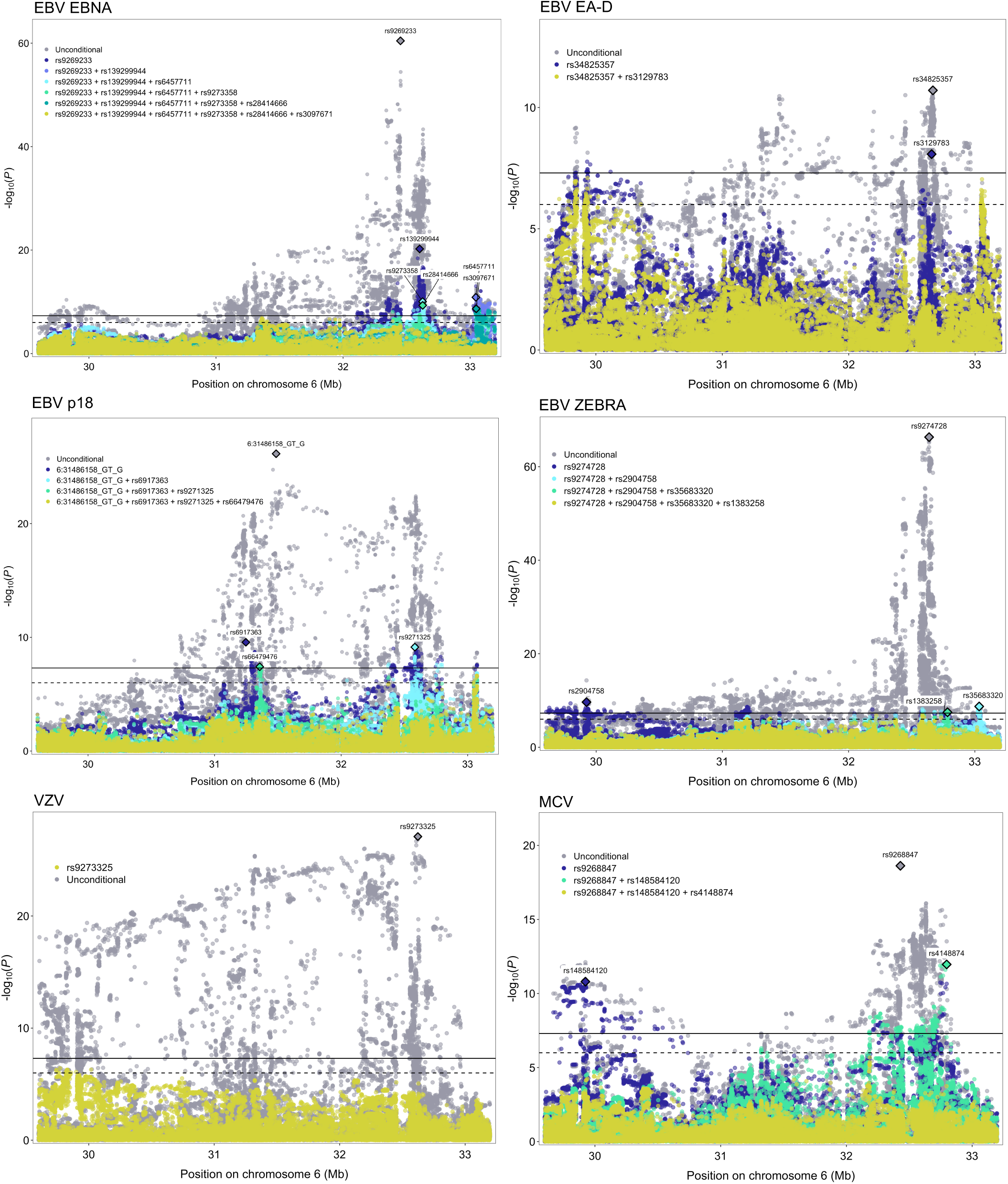
Regional association plots for conditionally independent HLA genetic variants that were significantly (*P*<5.0×10^-8^, solid black line) associated with each continuous antibody response phenotype. The suggestive significance threshold corresponds to *P*<1.0×10^-6^ (dotted black line).

Next, we tested 101 classical HLA alleles and performed analogous iterative conditional analyses for significantly associated variants (*P*<4.6×10^-5^). To help with the interpretation of our results, we depict the LD structure for HLA alleles in class II genes in Supplementary Figure 5. Significant associations across viruses were predominantly observed for class II HLA alleles. Five statistically independent signals were identified for antibody response to EBV ZEBRA (DRB4*00:00: β=-0.246, *P*=1.4×10^-46^; DQB1*04:02: β_cond_=0.504, *P*_cond_=1.0×10^-19^; DRB1*04:04: pcond=0.376, *P*_cond_=1.1×10^-18^; DQA1*02:01: β_cond_=0.187, *P*_cond_=1.1×10^-10^, A*03:01 : β_cond_=0.129, *P*_cond_=1.9×10^-8^) (**Figure 3; Supplementary Table 11**). DRB4*00:00 represents copy number absence, which co-occurs with DRB1*04 and DRB1*07 alleles^43^. This is consistent with the magnitude and direction of unconditional associations observed for DRB1*07:01 (β=0.251, *P*=1.3×10^-26^) and DRB4*04:01 (β=0.293, *P*=7.9×10^-22^). Five conditionally independent alleles were also identified for EBV EBNA: DRB5*00:00: β=-0.246, *P*=8.7×10^-30^; DRB3*02:02: β_cond_=0.276, *P*_cond_=6.8×10^-30^; DQB1*02:01: β_cond_=-0.164, *P*_cond_=3.6×10^-12^; DRB4*00:00: β=0.176, *P*_cond_=8.3×10^-17^; DPB1*03:01: β_cond_=-0.220, *P*_cond_=4.7×10^-14^ (**Figure 3; Supplementary Table 11**). DRB5*00:00 denotes a copy number deletion that sits on an common haplotype comprised of DRB1*15:01, DQB1*06:02, DQA1*01:02^43^, which may also include DRB5*01:01^44^ (**Supplementary Figure 6**). The presence of the DRB1*15:01-DQB1*06:02-DQA1*01:02 haplotype was associated with increased EBV EBNA seroreactivity (β=0.330, *P*=2.5×10^-28^). Fewer independent alleles were observed for EBV p18 (DRB5*00:00: β=-0.210, *P*=1.7×10^-22^; DRB1*04:04: β_cond_=0.357, *P*_cond_=1.3×10^-18^) (**Figure 3; Supplementary Tables 12**).

DQB1*02:01 was the only independently associated allele for EBV EA-D (β=-0.154, *P*=8.4×10^-11^) and HSV1 (β=0.145, *P*=2.8×10^-8^), although its effects were in opposite directions for each antigen (**Supplementary Table 13**). For VZV, associations with 16 classical alleles were accounted for by DRB1*03:01 (β=0.236, *P*=7.3×10^-26^). JCV shared the same lead allele as EBV EBNA and EBV p18 (DRB5*00:00: β=0.350, *P*=1.2×10^-21^) (Supplementary Table 13). Four conditionally independent signals were identified for MCV (DQA1*01:01: β=0.215, *P*=1.1×10^-15^; DRB1*04:04: β_cond_=-0.362, *P*_cond_=3.0×10^-11^; A*29:02: β_cond_=-0.350, P=1.0×10^-11^; DRB1*15:01: β_cond_=-0.203, *P*=3.7×10^-12^) (**Figure 3; Supplementary Table 14**). Lastly, we integrated associations across variant types by including conditionally independent HLA alleles as covariates in the SNP-based analysis. With the exception of EBV antigens and HHV7, classical HLA alleles captured all genome-wide significant SNP signals (**Supplementary Figure 7**).

Finally, we tested 980 HLA amino acid substitutions (**Supplementary Tables 15-24**), followed by omnibus haplotype tests at each position that had a significant amino acid and more than two possible alleles. The strongest allele-specific and haplotype associations were found at different positions in the same protein for EBV p18 (DRβ1 Ala −17: β=-0.194, *P*=1.0×10^-21^; DRβ1 (13): *P*_omni_=4.6×10^-22^), MCV (DQβ1 Leu-26: β=-0.173, *P*=7.0×10^-18^; DQβ1 (125): *P*_omni_=2.0×10^-17^), HHV7 (DQβ1 His-30: β=-0.111, *P*=1.2×10^-8^; DQβ1 (57): *P*_omni_=5.6×10^-9^), and HHV6 IE1B at (DRβ1 Ile-67: β=0.131, *P*=1.6×10^-8^; DRβ1 (13): *P*_omni_=1.1×10^-5^).

The strongest residue-specific and haplotype associations mapped to the same amino acid position for four phenotypes: EBV ZEBRA (**Supplementary Table 18**), HHV6 IE1A (**Supplementary Table 19**), HSV1 (**Supplementary Table 21**), and JCV (**Supplementary Table 23**). Amino acid residues at DQα1 (175) were associated with antibody response to EBV ZEBRA (Glu: β=0.279, *P*=1.1×10^-61^; *P*_omni_=8.3×10^-62^). Glu-175 is present in DQA1*02:01 (*P*=4.9×10^-27^), DQA1*03:01 (*P*=1.3×10^-16^), DQA1*04:01 (*P*=1.9×10^-12^), and seems to better summarize the EBV ZEBRA signal at this locus. Substitutions in DRβ1 (96) contained the strongest predictors of JCV seroreactivity (His or Tyr: β=0.325, *P*=1.6×10^-25^; *P*_omni_=7.7×10^-23^). His-96/Tyr-96 are in high LD (*r*^2^=0.92) with DRB5*00:00, the top JCV-associated allele. However, this might mask the signal for Gln-96 (β=-0.310, P=9.0×10^-23^), which is part of the DRB1*15:01 sequence (β=-0.309, *P*=9.0×10^-21^; LD *r*^2^=0.94). The lead signal for HSV1 mapped to DQβ1 (57) (Ala: (β=0.123, *P*=2.2×10^-10^; *P*_omni_=6.5×10^-9^), which aligns with the association for the lead HSV1-allele DQB1*02:01.

For EBV EBNA the strongest haplotype association was in DRβ1 (37) (*P*_omni_=1.1×10^-55^), while the residue with the lowest p-value was DQβ1 Ala-57 (β=-0.237, *P*=1.4×10^-42^) (Supplementary Table 16). Ala-57 maps to multiple DQB1 alleles and achieved a stronger signal for EBV EBNA than any classical HLA allele. Asp-9 in HLA-B showed the strongest association with antibody response to EBV EA-D (β=-0.146, P=1.8×10^-9^; Supplementary Table 15) and VZV (β=0.237, *P*=9.7×10^-25^; Supplementary Table 22). This amino acid sequence is part of B*08:01, which had analogous effects on both phenotypes (EBV EA-D: β=-0.144, P=2.7×10^-9^; VZV: β=0.238, *P*=4.7×10^-25^). Haplotypes with the lowest overall p-values were found in DQβ1 (71) for VZV (*P*_omni_=9.8×10^-19^) and DRβ1 (11) for EBV EA-D (*P*_omni_=1.7×10^-10^).

### TWAS of Genes Involved in Antibody Response

Based on known targets of infection or related pathologies, we considered expression in the frontal cortex (**Supplementary Table 25**), EBV-transformed lymphocytes for EBV antigens (**Supplementary Table 26**), and skin for MCV (**Supplementary Table 27**). Concordance across tissues was summarized using Venn diagrams (**Figure 4; Supplementary Figure 8**). TWAS identified 114 genes significantly associated (*P*_twas_<4.2×10^-6^) with antibody response in at least one tissue, 54 of which were associated with a single phenotype, while 60 influenced seroreactivity to multiple antigens. We also include results for 87 additional suggestively (*P*_twas_<4.2×10^-5^) associated genes.

The TWAS results included a predominance of associations in HLA class II genes. Some of the strongest overall associations were observed for *HLA-DRB5* (EBV ZEBRA: *P*_cortex_=4.2×10^-45^) and *HLA-DRB1* (EBV EBNA: *P*_cortex_=6.7×10^-39^; EBV ZEBRA: *P*_cortex_=3.3×10^-33^; JCV: *P*_cortex_=6.5×10^-14^; EBV p18: *P*_cortex_=2.2×10^-12^).

**Figure 4:**
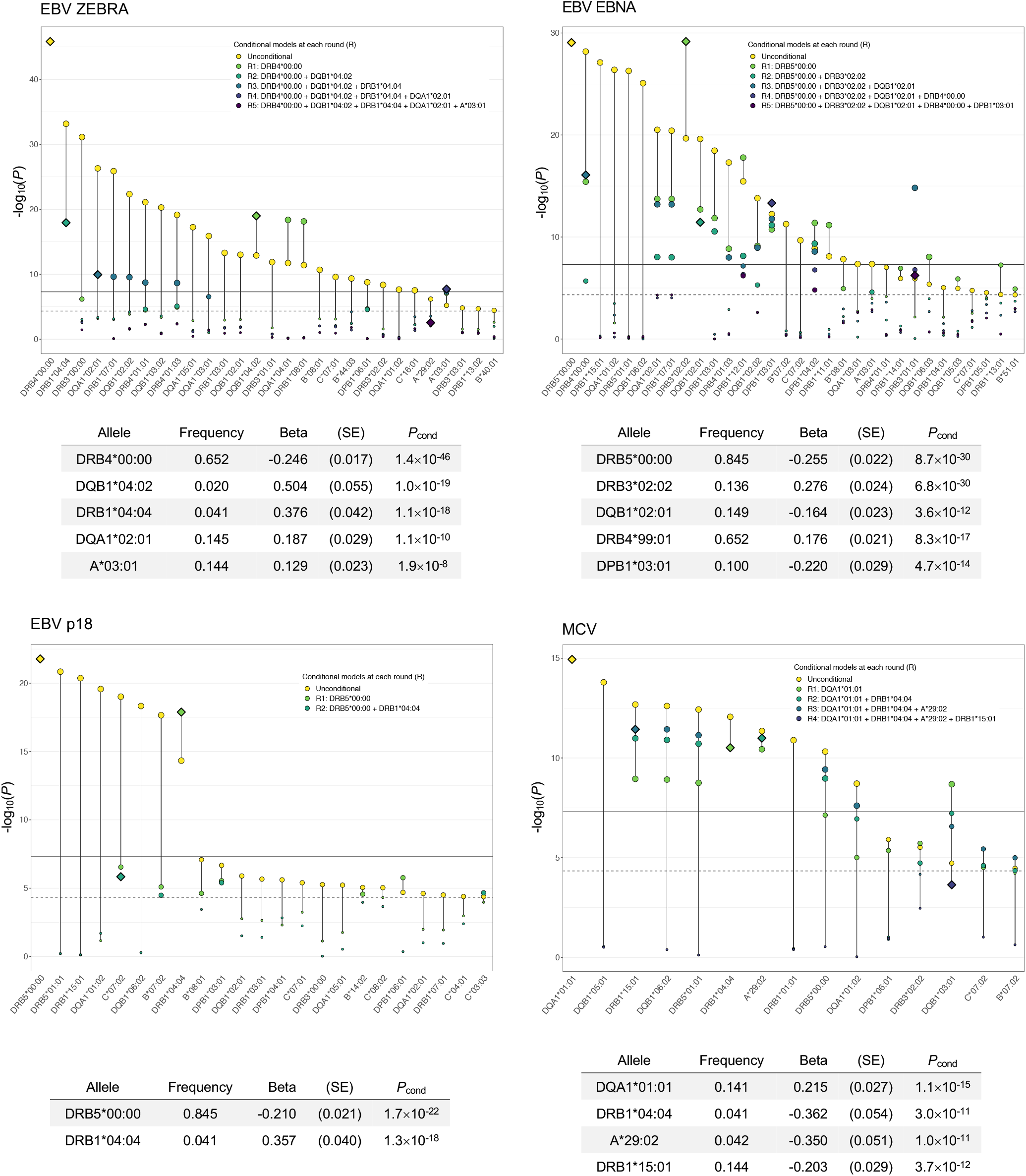
Conditionally independent classical HLA alleles significantly (*P*_cond_<5.0EBV EBNA10^-8^, solid line) associated with each continuous antibody response phenotype. Only classical alleles that surpassed the Bonferroni-corrected significance threshold (*P*<4.6×10^5^, dotted line) were included in conditional analyses.

**Figure 5:**
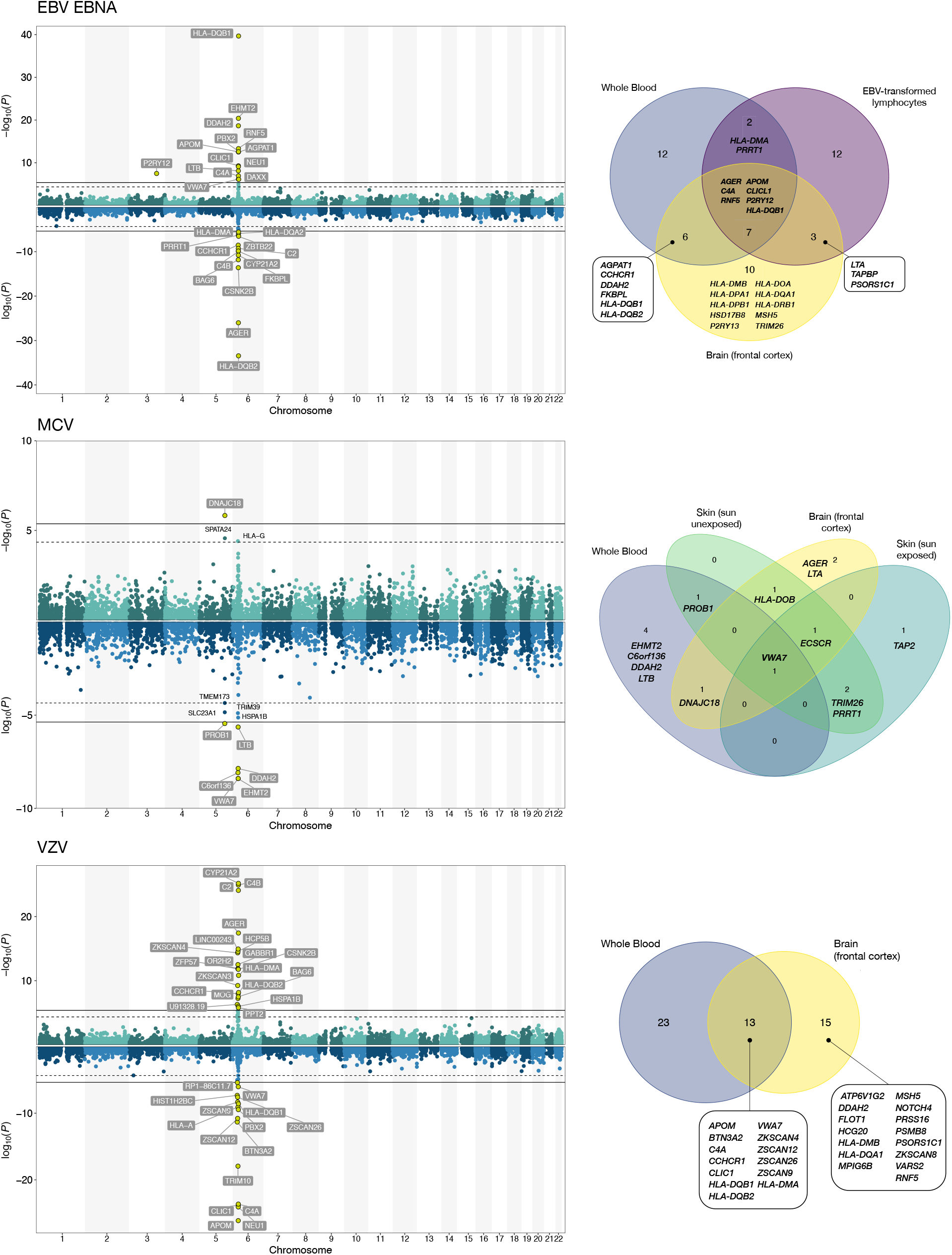
TWAS associations with continuous antigen response phenotypes. Two Manhattan plots depicting the transcriptome-wide associations for genes with a positive direction of effect (increased expression leads to higher antibody response) and genes with a negative direction of effect (increased expression is associated with a reduced antibody response).

Increased expression of *HLA-DQB2* was positively associated with antibody response to EBV ZEBRA (*P*_blood_=7.6×10^-19^), JCV (*P*_blood_=9.9×10^-10^), VZV (*P*_blood_=7.0×10^-9^), HHV7 (*P*_blood_=7.3×10^-8^), and HSV1 (*P*_blood_=3.3×10^-7^), but negatively associated with EBV EBNA (*P*_blood_=3.6×10^-34^) and EBV p18 (*P*_blood_=2.1×10^-8^), in a consistent manner across tissues. The opposite was observed for *HLA-DQB1*, with positive effects on EBV EBNA and EBV p18 and inverse associations with EBV ZEBRA, JCV, VZV, HHV7, and HSV1.

The TWAS analyses also identified a number of significant associations in the HLA class III region that were not detected in other analyses. The top-ranking VZV associated gene was *APOM* (*P*_blood_=7.5×10^-27^, *P*_cortex_=1.1×10^-25^). Interestingly, opposite directions of effect were observed for *C4A* and *C4B* gene expression. Increased *C4A* expression was positively associated with all EBV antigens (Supplementary Table 26), but negatively associated with VZV (*P*_blood_=2.3×10^-24^) and HSV1 (*P*_cortex_=1.8×10^-5^) antibody levels (Supplementary Table 25). On the other hand, increased *C4B* expression was inversely associated with EBV phenotypes, but positively associated with VZV (*P*_blood_=8.1×10^-25^) and HSV1 (*P*_blood_=1.1×10^-5^). A similar pattern was also observed for *CYP21A2* and *C2*, with positive effects on antibody response to VZV and HSV1, and negative effects for all EBV antigens. Other novel TWAS findings were detected for HHV7 in 22q13.2 *(CTA-223H9.9: P*_TWAS_=2.5×10^-6^; *CSDC2: P*_TWAS_=3.0×10^-6^; *TEF: P*_TWAS_=3.1×10^-6^) and 1q31.2 *(RGS1: P*_TWAS_=3.3×10^-6^).

The TWAS recapitulated several GWAS-identified loci: 3q25.1 for EBV EBNA *(P2RY13: P*_cortex_=1.1×10^-8^; *P2RY12:* Pbiood=3.3×10^-8^) and 19q13.33 for BKV *(FUT2: P*_TWAS_=8.1×10^-13^; *NTN5: P*_TWAS_=1.1×10^-9^). Transcriptomic profiles in skin tissues provided supporting evidence for the role of multiple genes in 5q31.2 in modulating MCV antibody response (Figure 4; Supplementary Table 27). The strongest signal was observed in for *ECSCR* (skin sun unexposed: *P*_TWAS_=5.0×10^-15^; skin sun exposed: *P*_TWAS_=4.2×10^-13^), followed by *PROB1* (sun unexposed: *P*_TWAS_=1.5×10^-11^). *ECSCR* expression was also associated based on expression in the frontal cortex, while *PROB1* exhibited a significant, but attenuated effect in whole blood. *VWA7* was the only gene associated across all four tissues for MCV and was also associated with antibody response to several EBV antigens.

Comparison of results for seroreactivity and seropositivity revealed a number of genes implicated in both steps of the infection process (**Supplementary Table 28**). Associations with HLA DQA and DQB genes in whole blood and HLA-DRB genes in the frontal cortex were observed for EBV antigens, JCV, and MCV. For MCV, the strongest seropositivity signals were observed for HLA class III genes *AGER* (*P*_cortex_=9.0×10^-21^) and *EHMT2* (*P*_blood_=5.8×10^-18^), which were also among the top-ranking genes for seroreactivity. Increased *ECSCR* expression conferred an increased susceptibility to MCV infection (*P*_cortex_=1.8×10^-8^), mirroring its effect on seroreactivity. In contrast to antibody response, no significant associations with any HLA genes were observed for VZV seropositivity.

Analyses using the Reactome database identified significant (q_FDR_<0.05) enrichment for TWAS-identified genes in pathways involved in initiating antiviral responses, such as MHC class II antigen presentation, TCR signaling, and interferon (IFN) signaling (**Supplementary Figure 9**). Pathways unique to herpesviruses included folding, assembly and peptide loading of class I MHC (q=3.2×10^-7^) and initial triggering of complement (q=9.8×10^-3^). Polyomaviruses were associated with the non-canonical nuclear factor (NF)-κB pathway activated by tumor necrosis factor (TNF) superfamily (q=1.9×10^-3^).

## DISCUSSION

We performed genome-wide and transcriptome-wide association studies for serological phenotypes for 16 common viruses in a well-characterized, population-based cohort. We discovered novel genetic determinants of viral antibody response beyond the HLA region for BKV, MCV, HHV7, EBV EBNA. Consistent with previous studies^7,8^ we detected strong signals for immune response to diverse viral antigens in the HLA region, with a predominance of associations observed for alleles and amino acids in *HLA-DRB1* and *HLA-DQB1*, as well as transcriptome-level associations for multiple class II and III HLA genes. Taken together, the findings of this work provide a resource for further understanding the complex interplay between viruses and the human genome, as well as a first step towards understanding genetic determinants of reactivity to common infections.

One of our main findings is the discovery of 5q31.2 as a susceptibility locus for MCV infection and MCV antibody response, implicating two main genes: *TMEM173* (or *STING1)* and *ECSCR*. The former encodes STING (stimulator of interferon genes), an endoplasmic reticulum (ER) protein that controls the transcription of host defense genes and plays a critical role in response to DNA and RNA viruses^45^. STING is activated by cyclic GMP-AMP synthase (cGAS), a cytosolic DNA sensor that mounts a response to invading pathogens by inducing IFN1 and NF-κB signalling^46,47^. Polyomaviruses penetrate the ER membrane during cell entry, a process that may be unique to this viral family^48^, which may trigger STING signaling in a distinct manner from other viruses^48^. Multiple cancer-causing viruses, such as KSHV, HBV, and HPV18, encode oncoproteins that disrupt cGAS-STING activity, which illustrates the evolutionary pressure on DNA tumor viruses to develop functions against this pathway and its importance in carcinogenesis^46^. Furthermore, cGAS-STING activation has been shown to trigger antitumor T-cell responses, a mechanism that can be leveraged by targeted immunotherapies ^49-51^. Several studies suggest STING agonists may be effective against tumors resistant to PD-1 blockade, as well as promising adjuvants in cancer vaccines^52-54^.

*ECSCR* expression in skin and brain tissues was associated with MCV antibody response and infection. This gene encodes an endothelial cell-specific chemotaxis regulator, which plays a role in angiogenesis and apoptosis^55^. *ECSCR* is a negative regulator of PI3K/Akt signaling by enhancing membrane localization of *PTEN* and operates in tandem with VEGFR-2 and other receptor tyrosine kinases^56^. In addition to 5q31.2, another novel MCV seroreactivity associated region was identified in 3p24.3, anchored by rs776170649, which has been linked to platelet phenotypes^57^. These findings align with a role of platelet activation in defense against infections via degranulation-mediated release of chemokines and β-defensin^58^.

Genetic variation within Fucosyltransferase 2 *(FUT2)* has been studied extensively in the context of human infections; however, its effect on BKV seroreactivity is novel. Homozygotes for the nonsense mutation (rs601338 G>A) that inactivates the FUT2 enzyme are unable to secrete ABO(H) histo-blood group antigens or express them on mucosal surfaces^59,60^. The allele which confers increased BKV antibody response (rs681343-T) is in LD (*r*^2^=1.00) with rs601338-A, the non-secretor allele, which confers resistance to norovirus^61,62^, rotavirus^63^, H. pylori^64^, childhood ear infection, mumps, and common colds^13^. However, increased susceptibility to other pathogens, such as meningococcus and pneumococcus^65^ has also been observed in non-secretors. Isolating the underlying mechanisms for BKV response is challenging because *FUT2* is a pleiotropic locus associated with diverse phenotypes, including autoimmune and inflammatory conditions^66,67^, serum lipids^68^, B vitamins^60,69^, alcohol consumption^70^, and even certain cancers^71^. In addition to *FUT2* in 19q 13.33, *NTN5* (netrin 5) suggests a possible link between BKV and neurological conditions. NTN5 is primarily expressed in neuroproliferative areas, suggesting a role in adult neurogenesis, which is dysregulated in glioblastoma and Alzheimer’s disease^72,73^.

We also report the first GWAS of serological phenotypes for HHV7. Genetic determinants of HHV7 antibody response in 6p21.32 were predominantly localized in *HLA-DQA1* and *HLA-DQB1*, with associations similar to other herpesviruses. In 11q23.3, rs75438046 maps to the 3’ UTR of *CXCR5*, which controls viral infection in B-cell follicles^74^, and *BCL9L*, a translocation target in acute lymphoblastic leukemia^75^ and transcriptional activator of the Wnt/β-catenin cancer signaling pathway^76^. In 17q21.32, *TBKBP1* encodes an adaptor protein that binds to TBK1 and is part of the TNF/NF-κB interaction network, where it regulates immune responses to infectious triggers, such as IFN1 signaling^77^. Interestingly, a protein interactome map recently revealed that SARS-CoV-2 nonstructural protein 13 (Nsp13) includes TBK1-TBKBP1 among its targets ^78^. Other functions of the TBK1-TBKBP1 axis relate to tumor growth and immunosuppression through induction of PD-L1^79^.

Several additional genes involved in HHV7 immune response were identified in TWAS. *TEF* in 22q13.2 is an apoptotic regulator of hematopoietic progenitors with tumor promoting effects mediated by inhibition of G1/S cell cycle transition and Akt/FOXO signaling^80^. *RGS1* in 1q31.2 has been linked to multiple autoimmune diseases, including multiple sclerosis^81^, as well as poor prognosis in melanoma and diffuse large B cell lymphoma mediated by inactivation of Akt/ERK^82,83^.

Other genes outside of the HLA region associated with viral infection response were detected for EBV EBNA in 3q25.1. The lead variant (rs67886110) is an eQTL for *MED12L* and *P2RY12* genes, which have been linked to neurodegenerative conditions^84,85^. *P2RY12* and *P2RY13*, identified in TWAS, are purinergic receptor genes that regulate microglia homeostasis and have been implicated in Alzheimer’s susceptibility via inflammatory and neurotrophic mechanisms^85^.

Considering genetic variation within the HLA region, our results confirm its pivotal role at the interface of host pathogen interactions and highlight the extensive sharing of HLA variants that mediate these interactions across virus families and antigens. Genes in this region code for cell-surface proteins that facilitate antigenic peptide presentation to immune cells that regulate responses to invading pathogens.

This region is critical for adaptive immune response but also has significant overlap with susceptibility alleles for autoimmune diseases. We identified 40 independent SNPs/indels associated with EBV (EBNA, EA-D, VCA p18, and ZEBRA), VZV, and MCV antibody response that accounted for all significant HLA associations for other phenotypes. Of the 14 conditionally independent, genome-wide significant classical alleles identified for 10 antigens, 7 were associated with multiple phenotypes. The most commonly shared HLA alleles were DRB5*00:00, DRB1*04:04, an known rheumatoid arthritis risk allele^86^, and DQB1*02:01, associated with celiac disease risk^87^. Copy number deletion represented by DRB5*00:00 may itself have a functional role in altering response by the absence of these alleles. DRB5*00:00 also summarizes signals from multiple HLA loci, including the extended DRB5*01:01-DRB1*15:01-DQB1*06:02-DQA1*01:02 haplotype that has been implicated in the etiology of multiple autoimmune diseases and EBV EBNA IgG levels. DRB1*15:01-DQB1*06:02-DQA1*01:02 is protective for type 1 diabetes^88^, while DRB5*01:01- DRB15:01 confers the strongest risk for developing multiple sclerosis^81^. Amino acid residues in DRβ1 at positions 11, 13, 71, and 74 and in DQβ1 codon 57 represent established susceptibility loci for rheumatoid arthritis^89^, type 1 diabetes^90^, and multiple sclerosis^91^ that exhibited strong associations with IgG levels for EBV, HHV7, VZV, JCV, and MCV antigens, and in some cases harbored the top signal of all HLA variants. Further research is needed to delineate shared genetic pathways that invoke autoimmunity and influence viral response.

Despite the predominance of association in HLA class II, several notable associations in HLA class I were detected. A*29:02 conferred reduced MCV seroreactivity and its sequence overlaps with amino acid residues in the A α1 domain (Thr-9, Leu-62, Gln-63, Asn-77, and Met-97) that were also significantly associated with decreased MCV antibody response. This is consistent with downregulation of MHC I as a potential mechanism through which Merkel cell tumors evade immune surveillance^92^. The strongest residue-specific signal for EBV EA-D and VZV mapped to B-Asp-9, which is located in the peptide binding groove and tags the B*08:01 allele, part of the HLA 8.1 ancestral haplotype. There is extensive evidence linking HLA 8.1, and B*08:01 specifically, with autoimmune diseases^93^ and certain cancers^94,95^, which may be attributed to its high cell-surface stability and increased probability of CD8+ T cell activation.

Comparison with other studies of host genetics and viral infection susceptibility shows that our results align with previously reported findings^7-9,96^ (**Supplementary Table 29**). We replicated most associations from two of the largest GWAS of humoral immune response in European ancestry subjects by Hammer et al.^7^ (n=2363) and Scepanovic et al.^8^ (n=1000), including HLA SNPs, alleles, amino acids, and haplotypes linked to EBV EBNA IgG, MCV IgG and serostatus, and JCV serostatus. We also replicated two *HLA-DRB1* variants (rs477515, rs2854275) associated with EBV EBNA antibody levels in a Mexican American population^9^. GWAS of HPV16 L1 replicated a variant previously linked to HPV8 seropositivity (rs9357152, *P*=0.008)^6^. Some of our findings contrast with Tian et al ^13^, although we confirmed selected associations, such as A*02:01 (shingles) with VZV (*P*=4.1×10^-8^) and rs2596465 (mononucleosis) with EBV EBNA (*P*=3.3×10^-9^) and EBV p18 (*P*=1.0×10^-12^). These differences may be partly accounted for by self-reported disease status in Tian et al. which is likely to reflect symptom severity and may be an imprecise indicator of infection with certain viruses or the magnitude of antibody response to infection.

One of the most striking findings in SNP-based HLA analyses was the genome-wide significant association between rs9273325, index VZV antibody response variant, and risk of schizophrenia. Previous epidemiologic and serologic studies have linked infections to schizophrenia, although the underlying mechanisms remain to be elucidated^97^. Viruses are plausible etiologic candidates for schizophrenia due to their ability to invade the central nervous system and disrupt neurodevelopmental processes by targeting specific neurons, as well as the potential for latent infection to negatively impact plasticity and neurogenesis via pro-inflammatory and aberrant immune signaling^97,98^. These observations are consistent with the established role the HLA region, including *HLA-DQB1*, in schizophrenia etiology^99,100^, and is further supported by previously reported associations for rs9273325 with blood cell traits^57^ and immunoglobulin A deficiency^101^, as well as its role as an eQTL for *HLA-DQB1* in CD4+ T2h cells. Schizophrenia susceptibility alleles DRB1*03:01^99^, DQB1*02:01, and B*08:01 were also the top three alleles associated with VZV antibody response in the unconditional analysis. Enhanced complement activity has been proposed as the mechanism mediating the synaptic loss and excessive pruning which is a hallmark of schizophrenia pathophysiology^102^. Complement component 4 (C4) alleles were found to increase risk of schizophrenia proportionally to their effect on increasing *C4A* expression in brain tissue^102^. Using gene expression models in whole blood and the frontal cortex we demonstrated that increased *C4A* expression is negatively associated with VZV antibody response. We also observed associations with *C4A* and *C4B* in EBV and HSV-1, but not other viruses. Taken together, these findings delineate a potential mechanism through which aberrant immune response to VZV infection, and potentially HSV-1 and EBV, may increase susceptibility to schizophrenia. However, cautious interpretation is warranted due to significant pleiotropy between HLA loci associated with viral infection and broad immune function.

Several limitations of this work should be noted. First, the UK Biobank is unrepresentative of the general UK population due to low participation resulting in healthy volunteer bias^103^. However, since the observed pattern of seroprevalence is consistent with previously published estimates^15^ we believe the impact of this bias is likely to be minimal on genetic associations with serological phenotypes. Second, our analyses were restricted to participants of European ancestry due to limited serology data for other ancestries, which limits the generalizability of our findings to diverse populations. Third, we were unable to conduct formal statistical replication of novel GWAS and TWAS signals in an independent sample due to the lack of such a population. Nevertheless, our successful replication of multiple previously reported variants and, combined with the observation that newly discovered genes and variants are part of essential adaptive and innate immunity pathways, support the credibility of our findings. Lastly, we also stress caution in the interpretation of GWAS results for non-ubiquitous pathogens, such as HBV, HCV, and HPV, due to a lack of information on exposure, as well as low numbers of seropositive individuals.

Our study also has distinct advantages. The large sample size of the UK Biobank facilitated more powerful genetic association analyses than previous studies, particularly in a population-based cohort unselected for disease status. Our detailed HLA analysis shows independent effects of specific HLA alleles and pleiotropic effects across multiple viruses. Analyses of genetic associations in external datasets further demonstrate a connection between host genetic factors influencing immune response to infection and susceptibility to cancers and neurological conditions.

The results of this work highlight widespread genetic pleiotropy between pathways involved in regulating humoral immune response to novel and common viruses, as well as complex diseases. The complex evolutionary relationship between viruses and humans is not dictated simply by infection and acute sickness, it is a complex nuanced architecture of initial challenge tempered with tolerance of viral latency over time. Yet it is that architecture that is evolutionarily optimized to maximize fitness early in life, the result of which may be increased risk for complex diseases later in life. Understanding this complex interplay through both targeted association studies and functional investigations between host genetic factors and immune response has implications for complex disease etiology and may facilitate the discovery of novel therapeutics in a wide range of diseases.

## Data Availability

The UK Biobank in an open access resource, available at https://www.ukbiobank.ac.uk/researchers/. This research was conducted with approved access to UK Biobank data under application number 14105 (PI: Witte).

## WEB RESOURCES

PLINK 2.0: https://www.cog-genomics.org/plink/2.0/

PLINK 1.07 conditional haplotype module: https://zzz.bwh.harvard.edu/plink/whap.shtml

R packages for pathway analysis: https://bioconductor.org/packages/release/bioc/html/ReactomePA.html and https://bioconductor.org/packages/release/bioc/html/clusterProfiler.html

Database of HLA allele frequencies and amino acid substitutions: http://www.allelefrequencies.net/hla9001a.asp?type_analysis=by_pops

## ACKNOWLEDGEMENTS

This research was supported by funding from the National Institutes of Health (US NCI R25T CA112355 and R01 CA201358; PI: Witte). Maike Morrison was funded by the University of California San Francisco’s Amgen Scholars Program.

